# Some principles for using epidemiologic study results to parameterize transmission models

**DOI:** 10.1101/2023.10.03.23296455

**Authors:** Keya Joshi, Rebecca Kahn, Christopher Boyer, Marc Lipsitch

## Abstract

**Background:** Infectious disease models, including individual based models (IBMs), can be used to inform public health response. For these models to be effective, accurate estimates of key parameters describing the natural history of infection and disease are needed. However, obtaining these parameter estimates from epidemiological studies is not always straightforward. We aim to 1) outline challenges to parameter estimation that arise due to common biases found in epidemiologic studies and 2) describe the conditions under which careful consideration in the design and analysis of the study could allow us to obtain a causal estimate of the parameter of interest. In this discussion we do not focus on issues of generalizability and transportability.

**Methods:** Using examples from the COVID-19 pandemic, we first identify different ways of parameterizing IBMs and describe ideal study designs to estimate these parameters. Given real-world limitations, we describe challenges in parameter estimation due to confounding and conditioning on a post-exposure observation. We then describe ideal study designs that can lead to unbiased parameter estimates. We finally discuss additional challenges in estimating progression probabilities and the consequences of these challenges.

**Results:** Causal estimation can only occur if we are able to accurately measure and control for all confounding variables that create non-causal associations between the exposure and outcome of interest, which is sometimes challenging given the nature of the variables we need to measure. In the absence of perfect control, non-causal parameter estimates should still be used, as sometimes they are the best available information we have.

**Conclusions:** Identifying which estimates from epidemiologic studies correspond to the quantities needed to parameterize disease models, and determining whether these parameters have causal interpretations, can inform future study designs and improve inferences from infectious disease models. Understanding the way in which biases can arise in parameter estimation can inform sensitivity analyses or help with interpretation of results if the magnitude and direction of the bias is understood.

## Introduction

In outbreaks of infectious diseases, models are often used to describe possible future scenarios. To use models most effectively for this purpose, it is important to have accurate estimates of parameters that describe the natural history of infection and disease in specific groups, such as the probability that an individual exposed to infection will experience various outcomes (e.g., infection, hospitalization or death) as well as related quantities such as the effectiveness of vaccination or prior infection in preventing infection or its consequences. Typically, estimates of relevant parameters are drawn from different studies with varying design and methods and starting at different points in the natural history of infection. During the COVID-19 pandemic, we have also seen that these parameters may be highly heterogeneous and can depend on a complex set of covariates; for example, the probability that an individual will be hospitalized if infected appears to depend on properties of the virus to which they are exposed (e.g. what variant), and properties of the individual such as their demographics (age and sex), comorbidities, and history of prior infection and vaccination. In this setting, it is important to delineate which estimands from empirical studies correspond to the quantities needed to parameterize transmission models.

We focus on parameterizing an individual-based model (IBM). The relationship between IBMs and epidemiologic study design has been only rarely discussed.^1,2^ An individual-based transmission model takes a population of individuals with given characteristics (e.g., age, sex, comorbidities, geographic location) and then simulates the process of transmission, whereby individuals are exposed to infection through their contacts with other infectious individuals, possibly while imposing interventions on other variables (e.g., vaccination). The consequences of infection, such as illness, hospitalization, or death, may then be explicitly tracked in the IBM. The resulting simulations can be used to help understand the effects of various countermeasures on the course of an outbreak.

In an individual-based model, each individual starts with a vector *L* of characteristics at baseline that are either fixed or follow their “natural course” throughout the simulation (for example, sex at birth and age respectively). We also include within *L* the comorbidities (e.g. diabetes) an individual may have at baseline that affect the risk of infection or downstream events given exposure. For the purposes of this discussion we include within *L* an individual’s history of prior infection (possibly with one or more of several variants) as a baseline covariate. During the simulation, other variables describing the individual may be altered through an intervention, such as vaccination. We call the vector of variables admitting interventions (other than exposure to the pathogen) *A*. Throughout this paper, to simplify the reasoning, we consider only interventions on a single variable, vaccination, and hereafter will use *A* and vaccination status interchangeably. At the start of the simulation we (for simplicity) set all but a small number of individuals to be unexposed (*E*=0) and typically introduce one or a few infectious individuals to initiate the transmission process. As the infection transmits, individuals may be exposed to infection, with a particular variant *e* of the infectious agent, setting their exposure status to *E=e*. To parameterize the IBM we need to estimate the probability of each of several consequences (defined below) for a person who has baseline characteristics *l*, has been assigned interventions *a*, and has been assigned exposure E=*e* with variant *e*. In the language of causal inference, this is the causal effect of jointly assigning vaccination *A*=1, and exposure *E*=*e* in a person with baseline characteristics *l*. For example, if H=1 is an indicator for hospitalization for COVID-19, an example parameter we might like to estimate is

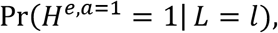

which can be read as the probability of hospitalization for COVID-19 for a person with baseline covariates *l* if they were given vaccination and exposed to SARS-CoV-2 variant *e*. In the COVID-19 context, there are several notable events downstream from exposure, shown in **Fig. 1A**.

**Figure 1:**
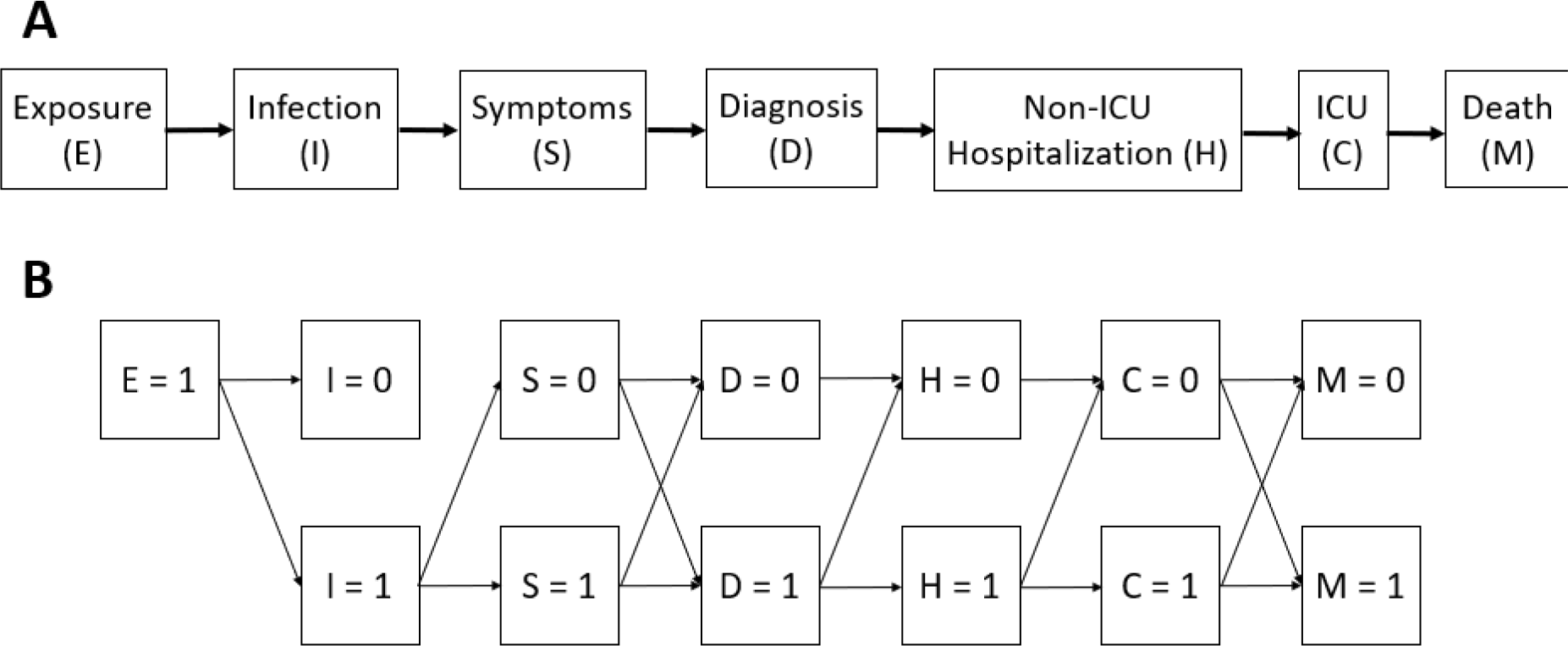
**(A)** Disease progression after exposure to COVID-19. (**B**) Combination of outcomes that can occur post exposure. The ordering described above is approximate in time, though some may coincide (e.g., hospitalization and diagnosis might be the same event) and some may be in the opposite order (e.g., diagnosis may precede symptom onset). This should not be taken as a compartmental model because the transitions are typically not Markov. That is, the probability of hospitalization given diagnosis will depend on symptoms (as well as on covariates).

In principle, if sufficient data were available, defining the probability of the outcome under this intervention is straightforward: for any person in the model exposed to infection, one would need the series of probabilities that the individual would experience each of the successive steps in the chain shown in **Figure 1A**, given *l* -- the individual’s demographic characteristics, immunocompromised status, comorbidities, and history of prior infection – if they are assigned to a specific vaccination status *a* and to be exposed to a particular variant *e*. If one knew the joint distribution of these probabilities from a study, one could draw from that distribution determined by *l* and *a* for each exposed individual at the time they were exposed and then progress the individual through the model states according to the drawn individual-specific outcome. Note that the probabilities of experiencing various outcomes might be correlated across individuals, both because some outcomes require previous ones (e.g., dying from infection can happen only if infected) and because individuals might vary in particular characteristics (e.g., immunodeficiency) that make them particularly likely to experience multiple outcomes (e.g., infection and hospitalization).

If the fate of a person (whether they experience downstream events after infection) is specified at the time of infection in the model, then the joint probabilities of each outcome will be used for parameterization, and the conditional probabilities of specific outcomes given previous ones will not be needed. However, depending on the details of how an IBM is parameterized, conditional probabilities may be required. If random draws are made for progression from each step (e.g., infection) to subsequent steps (e.g., hospitalization), then conditional probabilities will be needed. Though not the focus of this paper, we note that there may be good reasons to parameterize an IBM using joint or conditional probabilities or both, for example, to improve efficiency (which will often favor setting individuals’ fates at the start using joint distributions) or to accommodate time-varying interventions such as countermeasures (which will be far easier to parameterize if an individual’s use of the countermeasure can be decided at the time it is needed in the model). We focus on an IBM because there is a straightforward mapping of the quantities we want to estimate onto model parameters. For compartmental models we additionally have the problem that compartments will be heterogeneous, and thus even more approximations are needed.

To be more precise, let us restrict attention to six possible *events* that can occur after exposure: infection, symptoms, diagnosis, hospitalization, ICU, and death. In principle an individual set to vaccination status *a* and exposed to variant *e* progressing through the model will have one of 64 possible *outcomes*, with 0 (event did not occur) or 1 (event did occur) for each event. If we assume that each of these states were reachable only from the one before, then only 6 of the 64 possible outcomes are possible (0,0,0,0,0,0), (1,0,0,0,0,0), (1,1,0,0,0,0)…(1,1,1,1,1,1). In reality, some but not all of the states have this property. For example, infection = 1 is necessary for all downstream events to occur, but an individual may die without being admitted to the ICU. Therefore, the actual number of possible outcomes falls between 6 and 64, or more generally between *n* and 2^*n*^, where *n* is the number of events in the chain (**Figure 1B**). The probability of each of these possible outcomes may vary as a function of covariates including vaccination status, prior infection history, variant, and demographics and clinical risk factors.

Formally, the ideal would be, for each possible combination of outcomes {*I, S, D, H, C, M*}, to estimate Pr({*I, S, D, H, C, M*}^*e,a*^ | *L = l*) for all levels of exposure *e* and interventions *a* and where the vector L includes age, sex, immunocompromised status and comorbidities including prior SARS-CoV-2 infections (**Table 1**).

**Table 1:** Example covariates *L* that may predict the joint distribution of outcomes given infection.

| Name and meaning | Simple specification for COVID-19 | More detailed specification | Limitations of even the more detailed specification |
| --- | --- | --- | --- |
| Age | 5- or 10-year age group | 1 year age group | None |
| Sex | M, F, other | NA | None |
| Comorbidities and prior infection history | Prior infection and number of comorbidities from a list | Vector of presence/absence of individual comorbidities from a list and timing / number of prior infections | In practice data will be too sparse to estimate effects of all combinations of covariate presence/absence on the probabilities; may be incomplete in data source. Prior infection often unobserved |
| Immuno-compromised status | 0, 1 | Specific immunocompromising condition, neutrophil or CD4+ T cell count, etc. | Often measured with error |

Even with these simplifying assumptions, estimating all the parameters of interest in an IBM can be challenging for multiple reasons. First, it is typically impossible to know the joint distribution of all the states of interest from a particular study because we do not observe each of the successive outcomes for each person in the study. In particular, individuals experiencing more severe outcomes are more likely to be detected than those who experience milder outcomes and do not progress to severe disease.^3^ For example, we do not usually observe exposures or infections at all but only infer them from symptoms and/or diagnosis; we do not diagnose all infections but are more likely to do so if they are symptomatic, and even more if they are hospitalized. Second, in any practical study, the vector of patient and variant characteristics *L* will be high dimensional enough that nonparametric estimation will be infeasible and therefore we must use a parametric or semiparametric statistical model to estimate probabilities. For example, we might need to assume that variants and comorbidities act multiplicatively on an individual’s risk of an outcome because there are too few observations to estimate these relationships nonparametrically. Likewise, we often assume that vaccine effectiveness (VE) – the multiplicative reduction in risk for vaccinated vs. unvaccinated –is constant across demographic strata. Finally, there are various forms of bias that preclude accurate estimation of these quantities, even with unlimited data, including confounding and selection bias. This paper, using the example of COVID-19, describes these limitations and discusses different epidemiologic study designs that can help inform causal estimation of these parameters.

### Ideal design: human challenge trials where vaccination and exposure are randomized

We start by describing the ideal study design that would allow us to estimate all parameters of interest in the IBM under interventions on vaccination, noting this design, while possibly ethical in some situations,^4,5^ would be infeasible and unethical on the scale we are discussing,^6^ involving the need to observe severe and even fatal outcomes in multiple strata of *L*. In this hypothetical design, the investigator would take a simple random sample of individuals from the target population and then randomize them to both vaccination and exposure to a particular variant under conditions (e.g., infectious dose and setting) similar to those that would be encountered “naturally” outside the study.^7^ Participants would then be followed through the course of each postexposure outcome of interest allowing for the unbiased estimation of Pr({*I, S, D, H, C, M*}^*e,a*^|*L = l*) via observed probabilities Pr({*I, S, D, H, C, M*}|*E = e, A = a, L = l*).

Beyond ethical and feasibility concerns, other limitations of this design are that those that participate in the trial may be very different than those that do not (e.g. younger, healthier), preventing estimation of probabilities for many covariate strata, and it would not permit the estimation of the effects of vaccination on subsequent exposure, which may be important in parameterizing an IBM if awareness of receiving a vaccine leads to higher risk tolerance in social interactions.

### Alternative #1: vaccine trials and cohort studies to estimate ratios of marginal probabilities given exposure

When randomizing exposure is not feasible, one might instead consider using data from a randomized controlled trial (RCT) comparing vaccination versus placebo such as the phase III vaccine trials during the COVID-19 pandemic^8–10^. In most vaccine trials exposure is not directly measured, but it is possible to get unbiased estimates of marginal vaccine effectiveness (VE) on every downstream outcome. Under strong assumptions on homogeneity of effects, these VE estimates could then be combined with estimates of the (unvaccinated) probability of postexposure outcomes from contact-tracing studies in the same population to yield estimates of Pr({*I, S, D, H, C, M*}^*e,a*^|*L = l*) if we further assume that the covariates included in *L* are sufficiently rich that the population exposed to infection in the study is exchangeable with the target population of interest given L.

Under the assumption of no direct effects of vaccination on exposure, which should hold in a blinded placebo-controlled trial, vaccination status will be independent of exposure and thus we would expect equal rates of exposure in the vaccination groups in expectation. In this case, we can interpret the ratio of outcomes like symptomatic illness or hospitalization in these two groups as equal to the ratio of the probabilities of these outcomes conditional on exposure: exposure factors out of the ratio. Specifically, using conditional probabilities observable in the data, if we define *A =* 0 as being unvaccinated and *A =* 1 as being vaccinated,

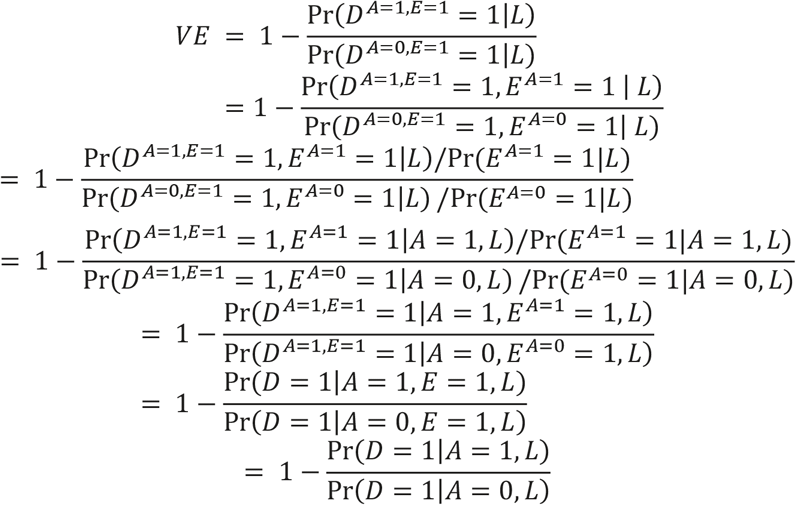

where the first equality is the definition of VE; the second recognizes that by definition *E =* 1 if *D =* 1, the third uses the fact that no effect of vaccination on exposure implies *E*^*A=*1^ *= E*^*A=*0^, the fourth uses conditional exchangeability (ensured by randomization), the fifth uses the definition of conditional probability, the sixth applies consistency and the last again follows from the fact that *E =* 1 if *D =* 1. Thus, while we cannot get absolute estimates of the probabilities, we can ascertain multiplicative effects of vaccination.

Unfortunately, large vaccine trials are often expensive and logistically difficult. Therefore, relevant trial data may not be available for all postexposure outcomes (particularly rare ones) and, over time, the exposure conditions may be invalidated by the emergence of new variants. In this case, one may instead attempt to emulate a vaccine trial using observational data. Examples in COVID-19 include matched cohort studies using electronic healthcare records^11^. In these studies neither exchangeability between the vaccinated and unvaccinated in their counterfactual outcomes given exposure, nor exchangeability in their exposure, is ensured by design, but rather must be assumed to be approximately true conditional on *L*. If this assumption is correct, observational emulations can also be used to estimate VE under the same arguments above about the expected equivalence of exposure and counterfactual outcomes given exposure within strata defined by *L*. However, in the absence of blinding, excluding effects of vaccination on exposure may be less plausible.

Other observational study designs are also commonly used to estimate vaccine effectiveness. Notably, the test-negative design in which individuals presenting at a given level of severity with and without a positive test for SARS-CoV-2 (symptomatic infection or hospitalization, most commonly) are compared with respect to their prior vaccination status. Much has been written about the advantages and limitations of these designs for estimating vaccine effectiveness.^12–14^ Without recapitulating that discussion, we note that in these studies, vaccine effectiveness is defined as one minus the odds ratio of infection for vaccinated vs. unvaccinated persons, and if other conditions for validity are met, these studies too can provide estimates of the causal effect of vaccination on the risk of an outcome given exposure.^14,15^

Note that these studies provide useful information for studying the effects of vaccination but do not so readily inform questions related to viral characteristics,^16^ including variants, since it is implausible to assume exposure to a given variant (e.g., delta and omicron) is equal on any given day, conditional on an individual’s covariates. However, a cohort study or randomized trial performed over a period of time spanning the periods of predominance of two or more variants could in principle estimate vaccine effects separately in the different periods and compare these. In practice, we are unaware of such comparisons in cohort or randomized studies, though they have been performed in other observational study designs.^17^

### Alternative #2: contact-tracing studies to estimate marginal and joint conditional probabilities of outcomes given exposure and covariates

In some cases, such as in the immediate aftermath of the emergence of a new variant, one might consider following contacts of confirmed cases to estimate Pr({*I, S, D, H, C, M*}^*e,a*^|*L = l*). Such studies have been performed and are typically based on contact tracing. These studies identify household^18^ or community ^19,20^ contacts of known cases and follow their outcomes. In this design, which is an observational analog of the challenge trial described above, causal effects of vaccination can be estimated under the strong assumptions that both selection on exposure and vaccination are exchangeable conditional on the covariates in *L* and, again, that there are no effects of vaccination on exposure.

While these types of studies collect detailed information on individuals, they are often small because of the effort required to identify and follow contacts. Nonetheless, large contact-tracing-based studies are possible and have provided valuable estimates of some of these probabilities as well as of additional parameters beyond those considered here, such as vaccine effects on infectiousness.^21^ On the other hand contact-tracing studies, through closer follow up with participants, often permit more detailed data collection than is possible in a large trial or cohort study. In particular, they can be useful for closely documenting intermediate disease states (such as asymptomatic infection^19^) and biomarkers such as correlates of protection and viral load that may be missing in larger studies.

### Progression (conditional) probabilities: Caution warranted

As noted in the introduction, we may wish to use progression probabilities: the probability of moving from one state to the next in Fig. 1, to parameterize an individual-based model. However, in most cases, estimating progression probabilities is not straightforward, and there are more potential sources of bias. One can design a study specifically to estimate these progression probabilities. For logistical reasons, it is often convenient to follow individuals who are ascertained at an event in Fig. 1 later than exposure and estimate probabilities of subsequent events conditional on the ascertaining event. For example, studies may ascertain individuals at the time of symptoms or diagnosis^22^ and assess the relative probability of hospitalization or death conditional on that ascertainment event, among different groups of individuals defined by covariates. Similarly, some studies ascertain individuals at the time of hospitalization^23^ and assess their clinical course and risk of mortality. In what follows we describe issues in parameter estimation arising from such studies, using the exposure of vaccination as an example.

One major threat for accurate VE estimates for model parameterization is conditioning on a post-exposure variable. For example, vaccine effectiveness against progression to symptoms (VE_P_) is an estimate among those who are infected, not among all exposed individuals.

As another example which we consider in more detail, suppose one wanted to estimate the impact of vaccine in preventing hospitalization among individuals with symptomatic infection, a form of VE_P_ where the progression is from symptomatic infection to hospitalization. We define VE_S_ as vaccine effectiveness against symptomatic infection; VE_SH_ as vaccine effectiveness against hospitalization, a function of protection against symptomatic infection and hospitalization; and VE_H|S_ as vaccine effectiveness against hospitalization among individuals who are symptomatically infected.^24^ Defining each type *j* ∈ {*S, SH, H*|*S*} of *VE*_*j*_ we can obtain:

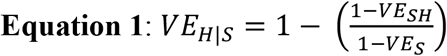

Given that some parameterizations of an IBM might require a conditional measure such as *VE*_*H*|*S*_ it is important to understand what estimate of this quantity we can obtain from a study, and if that parameter has a causal interpretation.

Fig.2 shows the directed acyclic graph (DAG) describing some of variables that influence the probability of vaccination and all downstream events. By conditioning on symptomatic infection, we create a selection bias and induce a non-causal association between hospitalization and vaccination because symptom status is a collider (e.g., a common effect of two variables). As a result of this selection bias, the vaccine may incorrectly appear to increase the risk of hospitalization, or its protective effect may be estimated with bias. To minimize the impact of this selection bias, we need to adjust for all variables that confound (e.g., a variable that, if conditioned upon, will close all non-causal paths between the exposure and outcome of interest) the relationship between hospitalization and symptomatic infection.

**Figure 2:**
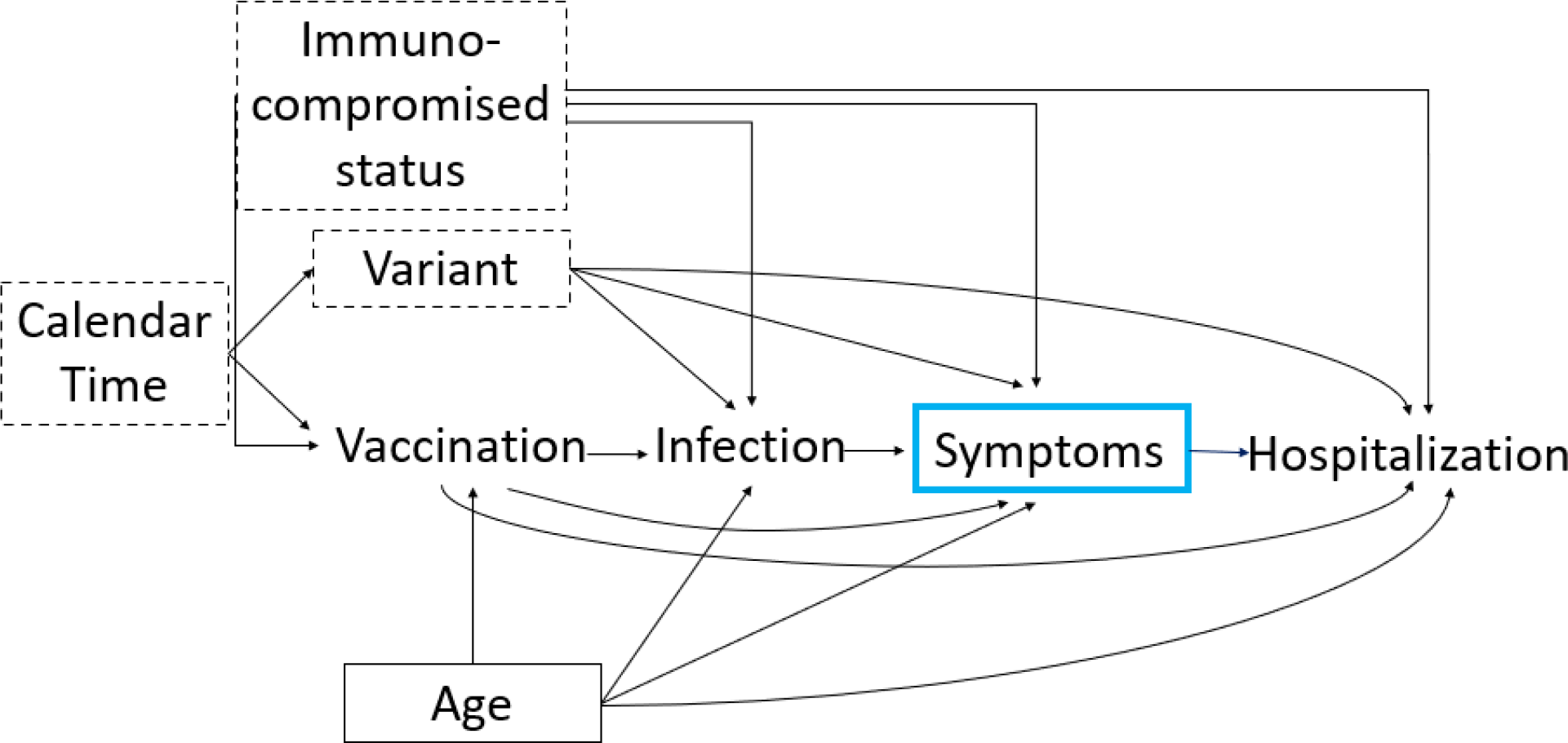
Distortion of VE_H|S_ estimates due to selection bias induced by conditioning on a collider, symptomatic infection. Directed acyclic graph (DAG) showing variables that can impact the relationship between vaccination and hospitalization among individuals with symptomatic disease. Blue box means a variable is conditioned upon. Variables with a solid box around them can be fully controlled for using stratification-based approaches. Variables with dashed lines can be controlled for, but likely with some error resulting in residual confounding.

Some confounders, including age, can be measured, and adjusted for by stratification or modeling. However, there are other confounders that can be measured with varying degrees of accuracy including calendar time and variant (Fig 2). In the absence of universal sequencing, accurately ascertaining which variant an individual is infected with is challenging due to co-circulation of variants over time. Previous studies^22^ have shown that certain proxies, including S-gene target failure (SGTF) can be used to accurately identify which variant an induvial has been infected with. If neither is available, misclassification of the infecting variant is possible, and can bias effect estimates of disease severity^25^ and VE as we are not fully able to adjust for the impact of variant through stratification-based approaches. Calendar time could be controlled for in a cohort study where one could match exposure groups on this variable (i.e., day). For other designs, including case-control studies, adequately controlling for this variable could be challenging.

In addition, immunocompromised status is challenging to measure as it is influenced by multiple factors (e.g., genetics, environmental, clinical, and demographic characteristics), and will likely be misclassified; in particular, immunocompromising conditions placing their carriers at risk for severe outcomes of COVID-19 infection may be undocumented at baseline in many individuals.^26^ If we cannot control for these variables using stratification, VE estimates will be biased. Thus, if the full set of variables, in this example, age, variant, and immunocompromised status cannot be adjusted for either in the design or analysis phase of the study, then the effect estimate will be biased, and there will be no causal interpretation. Therefore, if it is possible to measure confounding variables accurately, design and analysis of studies should prioritize collection and adjustment of these variables.

More likely, it will not be possible to accurately measure the variables needed to close all non-causal paths between vaccination and hospitalization among those with symptomatic infection, or data on these variables will be unavailable (e.g., if using pre-existing data or data collected for other purposes).^18^ If some data are available, proxies or cruder classifications of the covariates of interest (e.g., age bins vs age in years) could be used. In this scenario there will be residual confounding and the parameter of interest will have some bias, however it is an estimate that could still be used in the study population in the absence of higher quality information.

Even if we were able to control for all confounders, the estimate of conditional vaccine effectiveness sill has an ambiguous causal interpretation as the set of individuals who would become infected with symptoms if vaccinated is likely not identical to the set that would be symptomatically infected given no-vaccination.^24,27^ Methods to adjust for post-treatment variables have previously been developed and applied to study causal vaccine effects in rotavirus and pertussis.^27^ Under strong assumptions, the causal effect for certain subgroups, those who would always get infected irrespective of vaccination status, can be identified.^24,27^ While this subgroup parameter will have a casual interpretation, the population this estimate can be generalized to is limited and how to apply it in an IBM may not be straightforward.

Sensitivity analyses for the effects of unmeasured confounding can help to assess the degree of plausible bias, as has been illustrated in a related context, studying the impact of variant on the probability of hospitalization given diagnosis^22^ and accounting for potential downward bias in the estimate of Omicron (vs. Delta) per-case hospitalization risk due to an association between being an Omicron case and higher prevalence of prior infection.^25^

A second common approach for measuring progression probabilities is using a disease severity pyramid.^28–30^ Comparing the probability of a severe outcome (e.g., death or hospitalization) given that an individual was infected (at the population level this is the infection-fatality ratio) or symptomatic (at the population level this is the symptomatic case-fatality ratio) between groups that have received different interventions is crucial in epidemiologic investigations of infectious disease outbreaks.^31^ To estimate the symptomatic case fatality ratio (sCFR), one could use the conditional probabilities of surviving up to the state immediately preceding. Thus, the sCFR could be decomposed into:

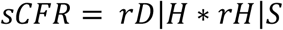

where rD|H is the probability of death given hospitalization and rH|S is the probability of hospitalization given symptomatic infection.

In addition to the challenges mentioned above, “severity pyramid” analyses often synthesize multiple data sources where a given study provides information about a specific level of the pyramid. This analysis therefore assumes that an individual who makes it through a level in a given study is exchangeable with an individual that starts at the next level in a different study. This is a strong assumption that might not be realistic in practice due to both selection bias and confounding.

#### Interpretation challenges

Conditional estimates can also pose interpretation challenges. One might expect that VE_H|S_ should only decrease with time since vaccination, due to waning vaccine effectiveness. However, these estimates can either increase or decrease with time since vaccination. This phenomenon occurs because VE_H|S_ is a composite measure of two values of VE that may wane at different rates.^32^ In **Table 2**, we show two examples in which both VE_S_ and VE_SH_ wane over time, but the trends for VE_H|S_ differ. In Example 1, VE_H|S_ is lower for those vaccinated longer ago than for those more recently vaccinated, as might be naively expected in the presence of waning vaccine-derived immunity. In Example 2, protection against infection, VE_S_, wanes at a much faster rate than protection against hospitalization, VE_SH_, and VE against hospitalization conditioned on infection (VE_H|S_) increases with time since vaccination. These trends are the result of defining an “effectiveness” measure conditional on the post-treatment variable of infection, which mediates part of the protection a vaccine offers against hospitalization. Therefore, while a conditional measure like VE_H|S_ may be useful for model parameterization, these subtleties in its interpretation lead us to question its usefulness, in contrast to the arguments of ref.^33^

**Table 2:** Example where VE_S_ and VE_SH_ wane at similar rates with effectiveness estimates similar to mRNA-vaccine effectiveness for Delta variant (Example 1) vs an example where VE_S_ wanes much faster than VE_SH_ as seen for mRNA-vaccine effectiveness estimates for Omicron variant (Example 2).

| Vaccination | Example 1<br>$VE_S$ | Example 1<br>$VE_{H\vert S}$ | Example 1<br>$VE_{SH}$ | Example 2<br>$VE_S$ | Example 2<br>$VE_{H\vert S}$ | Example 2<br>$VE_{SH}$ |
| --- | --- | --- | --- | --- | --- | --- |
| **Recent** | 90% | 70% | 97% | 62% | 63% | 86% |
| **Longer ago** | 65% | 43% | 80% | 0% | 71% | 71% |

As noted earlier, parameterization of the IBM can happen in multiple ways. If conditional measures are estimated or are needed for parameterization, analysts should keep in mind that if two forms of effectiveness wane at different rates, as has been suggested throughout the COVID-19 pandemic, their ratio will not necessarily wane over time. Indeed, it would be ideal not to refer to VE_H|S_ as effectiveness at all, though it may be too late to change the terminology.

## Discussion

We have discussed approaches to parameter estimation for IBMs and challenges arising due to common biases found in epidemiologic studies using relevant examples from the COVID-19 pandemic. Ideal designs, which would allow estimation of all parameters of interest in the IBM by randomizing exposure, including human challenge trials, might not be ethical or feasible, or if they were, would provide limited insight on the effect of vaccination on downstream events in key groups, including older adults and individuals with comorbidities.

If exposure cannot be randomized, one can get valid estimates of marginal VE on downstream outcomes by balancing exposure between vaccinated and unvaccinated individuals in a RCT or matched cohort studies under assumptions about conditional exchangeability given L. From these designs one can only estimate multiplicative effects of vaccination, not absolute estimates of the probabilities of interest. In addition, these designs are not suitable for understanding how the risk of infection and subsequent disease progression changes across variants, which can more likely be answered using contact tracing based study designs.

In many instances we may be interested in the probability of moving from one state to the next in an IBM. Estimating these progression probabilities can be challenging and are subject to multiple sources of bias including confounding and selection bias. Depending on the type of confounding variable, full control is likely impossible. Even if control of all confounders was achievable, the progression probability does not have a precise casual interpretation.

However, even in the absence of perfect control, non-causal parameter estimates may still be used, as sometimes they are the best available information we have. The potential sources of bias and ways to mitigate them that we have outlined here can be used to help inform the design and analysis of future studies. Understanding the way in which biases can arise in parameter estimation can inform sensitivity analyses or help with interpretation of results if the magnitude and direction of the bias is understood.

## Data Availability

No code was used in this paper.

## Acknowledgements

We thank Dr. Jason Asher for his helpful discussions.

